# Efficacy of remdesivir-containing therapy in hospitalized COVID-19 patients: a prospective clinical experience

**DOI:** 10.1101/2021.07.01.21259852

**Authors:** Alessandro Russo, Erica Binetti, Cristian Borrazzo, Elio Gentilini Cacciola, Luigi Battistini, Giancarlo Ceccarelli, Claudio Maria Mastroianni, Gabriella d’Ettorre

**Author notes:** **Corresponding author** Prof. Alessandro Russo, Infectious and Tropical Disease Unit, Department of Medical and Surgical Sciences, Viale Europa, 88100, “Magna Graecia” University of Catanzaro, Italy.

## Abstract

**Objectives:** remdesivir is currently approved for the treatment of COVID-19. The recommendation for using remdesivir in COVID-19 was based on the *in vitro* and *in vivo* activity of this drug against SARS-CoV-2.

**Methods:** this was a prospective, observational study conducted on a large population of patients hospitalized for COVID-19. The primary endpoint of the study was to evaluate the impact of remdesivir-containing therapy on 30-day mortality; secondary endpoint was the impact of remdesivir-containing therapy on the need of high flow oxygen therapy (HFNC) or non-invasive ventilation (NIV) or mechanical ventilation. Data were analyzed after propensity score matching.

**Results:** 407 patients with SARS-CoV-2 pneumonia were consecutively enrolled. Out of these, 294 (72.2%) and 113 (27.8%) were respectively treated or not with remdesivir. Overall, 61 (14.9%) patients were treated during hospitalization with non-invasive or mechanical ventilation, while a 30-day mortality was observed in 21 (5.2%) patients with a global in-hospital mortality of 11%. Cox regression analysis, after propensity score matching, showed that therapies, including remdesivir-containing therapy, were not statistically associated with 30-day survival or mortality, while need of HFNC/NIV (HR 17.921, CI95% 0.954-336.73, p=0.044) and mechanical ventilation (HR 3.9, CI95% 5.36-16.2, p=0.003) resulted independently associated with 30-day mortality. Finally, therapies including or not remdesivir were not independently associated with a lower or higher risk of HFNC/NIV or mechanical ventilation.

**Conclusions:** this real-life experience about the remdesivir use in hospitalized patients with COVID-19 was not associated with significant increase in rates of survival or reduced use of HFNC/NIV or mechanical ventilation, compared to patients treated with other therapies not including remdesivir.

## INTRODUCTION

Severe acute respiratory syndrome coronavirus-2 (SARS-CoV-2) is identified as the cause of an outbreak of respiratory illness, evolving in a pandemic. The virus causes respiratory illness and can rapidly spread from person to person; then, in a large number of patients this virus causes coronavirus diseases (COVID-19) characterized by pneumonia, severe acute respiratory syndrome, kidney failure, and a significant rate of mortality [1-2-3].

To date, treatment of critically ill infected patients is primarily supportive with a robust evidence reported in literature about the use of steroids, especially dexamethasone, in lowering mortality especially in critically ill patients [4]; some important evidence suggests also the role of low-molecular-weight heparin (LMWH) to reduce in-hospital mortality [5].

As a matter of fact, only remdesivir was currently approved for the treatment of COVID-19. The recommendation for using remdesivir as treatment of COVID-19 is based on the *in vitro* and *in vivo* activity of remdesivir against SARS-CoV-2 [6]. Moreover, remdesivir showed an acceptable safety profile and exhibits *in vivo* prophylactic and therapeutic efficacy against SARS-CoV-2 infection [7].

According to randomized clinical trials, its administration might shorten the time to recovery and reduce severity of infection in adults hospitalized with COVID-19 [8]. However, there are still many ongoing clinical trials and more evidence is needed to confirm the efficacy of remdesivir in treating patients with COVID-19 at different stages of severity.

Aim of our study was to evaluate in a real-life, prospective, experience the impact of remdesivir on 30-day mortality and the need of non-invasive or invasive ventilation in hospitalized patients with SARS-CoV-2 pneumonia.

## RESULTS

During the study period 407 patients with SARS-CoV-2 pneumonia were consecutively enrolled. Out of these, 294 (72.2%) and 113 (27.8%) were respectively treated or not with remdesivir. Overall, 61 (14.9%) patients were treated during hospitalization with non-invasive or mechanical ventilation, while a 30-day mortality was observed in 21 (5.2%) patients, with a global in-hospital mortality of 11%.

In **Table 1** is reported univariate analysis about demographics and clinical characteristics of COVID-19 patients treated or not with remdesivir. Statistically significant differences were observed in remdesivir group about male sex (80% Vs 62%, p<0.001), fever (79% Vs 50%, p<0.001), cough (50% Vs 29%, p<0.001), dyspnea (57% Vs 37%, p<0.001), compared to patients not treated with remdesivir. No statistically significant differences were observed in the remdesivir group about age (63.2 Vs 62.5 years, p=0.717), length of hospital stay (15.02 Vs 16.06 days, p=0.487), and days to nasopharyngeal swab negativization (22.07 vs 24.77 days, p=0.378). Finally, no differences were observed about bacterial co-infection (20% Vs 21%, p=0.928) and 30-day mortality (4% Vs 6%, p=0.411).

**Table 1.**
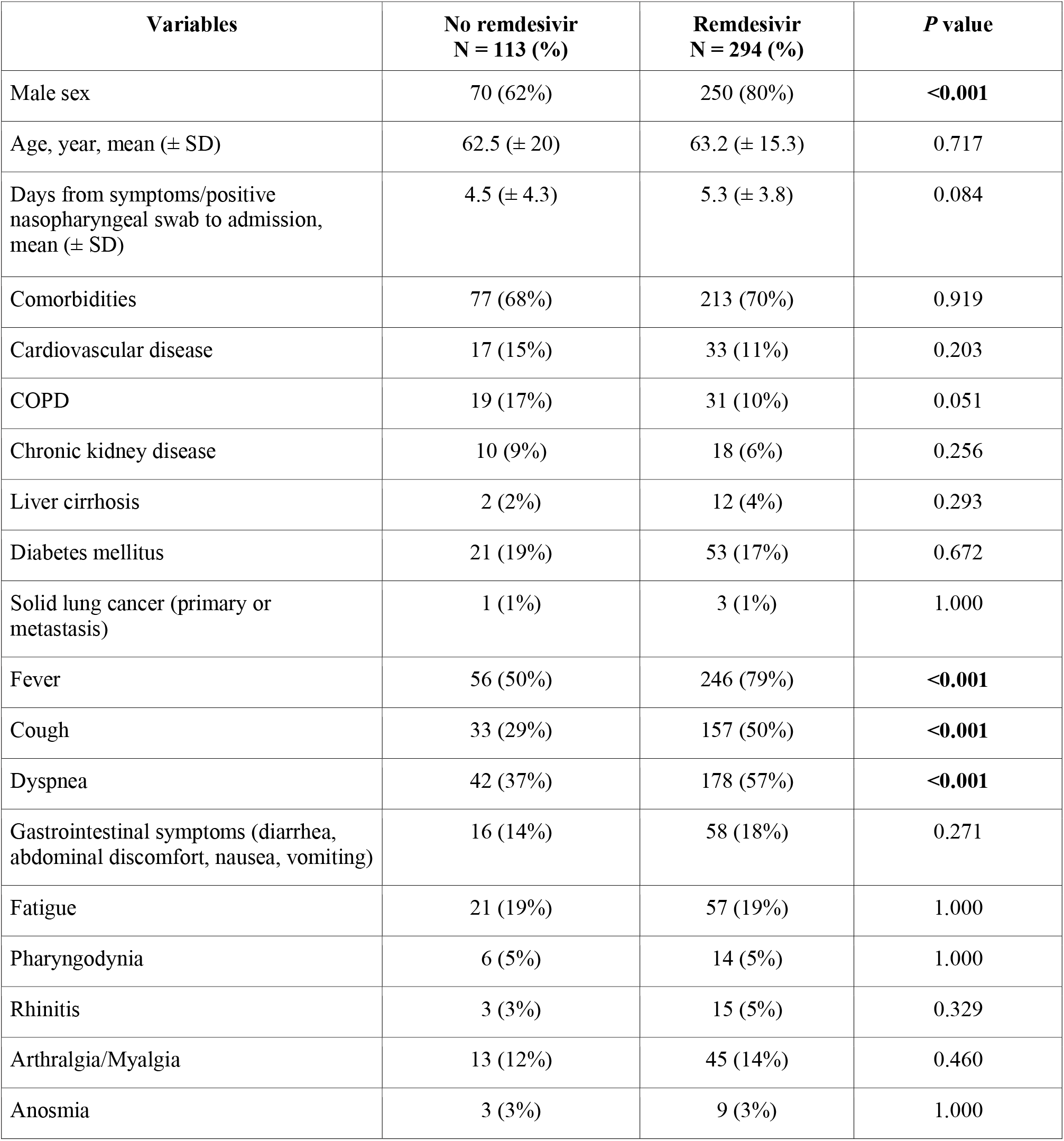
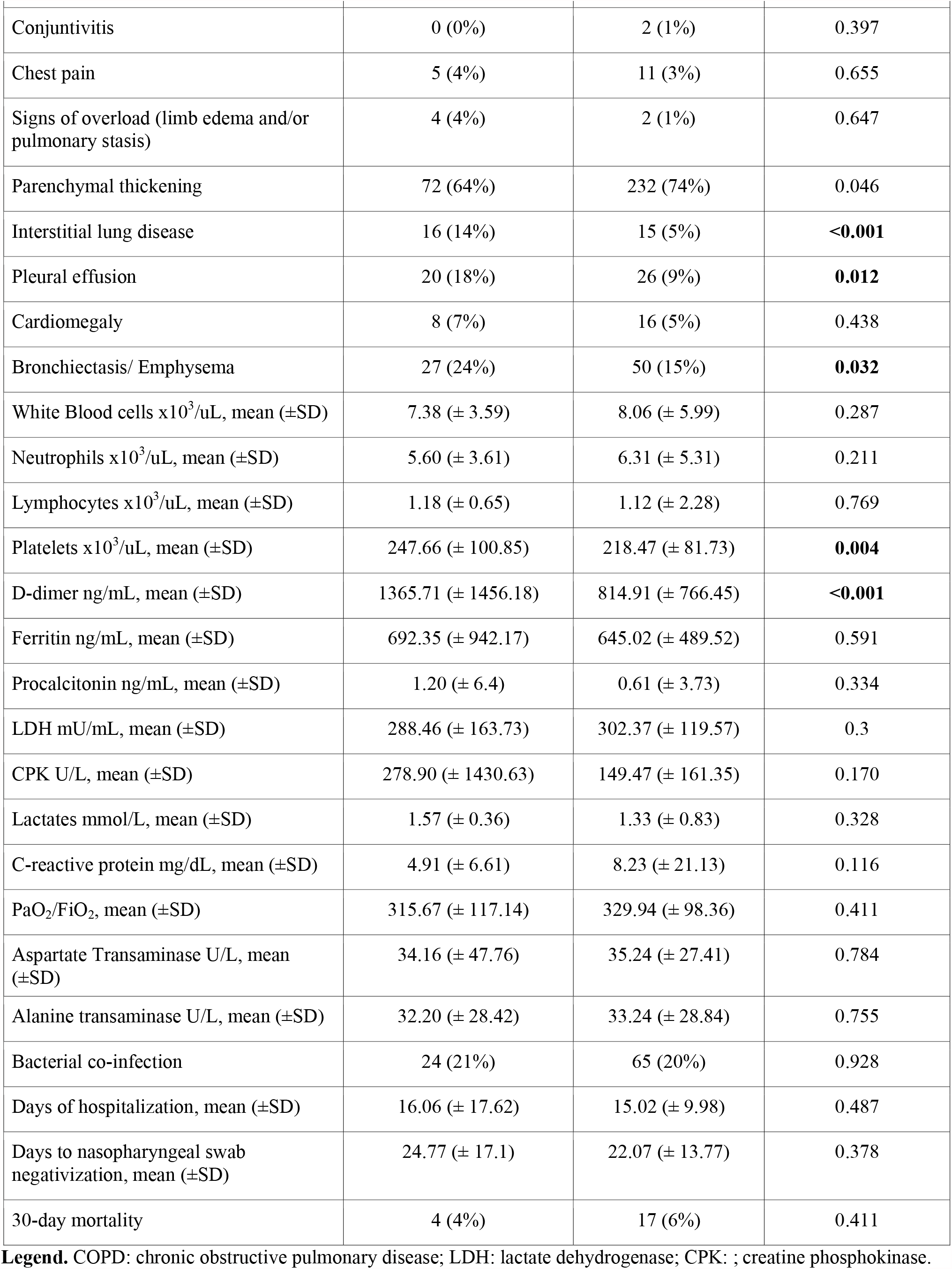
Univariate analysis about demographics and clinical characteristics of COVID-19 patients treated or not with remdesivir

Other treatments used in COVID-19 patients were reported in **Table 2**. Comparison between patients treated or not with remdesivir showed that steroids (93% Vs 81%, p<0.001), LMWH (93% Vs 52%, p<0.001) were more frequently prescribed in remdesivir group. Antibiotic therapy (58% Vs 27%, p<0.001) was more frequently prescribed for patients not treated with remdesivir, while no differences were reported about the use of HFNC/NIV or mechanical ventilation in the 2 study groups.

**Table 2.** Other treatments in COVID-19 patients treated or not with remdesivir

| Variables | No remdesivir<br>N = 113 (%) | Remdesivir<br>N = 294 (%) | *P* value |
| --- | --- | --- | --- |
| Steroids | 92 (81%) | 289 (93%) | **<0.001** |
| Antibiotics (excluding macrolides) | 65 (58%) | 83 (27%) | **<0.001** |
| Macrolides | 74 (65%) | 146 (46%) | **<0.001** |
| Low-molecular-weight heparin | 59 (52%) | 280 (93%) | **<0.001** |
| Low-flow oxygen therapy / no need of oxygen therapy | 89 (79%) | 275 (87%) | **0.044** |
| HFNC/NIV | 17 (15%) | 33 (10%) | 0.155 |
| Mechanical ventilation | 1 (1%) | 10 (3%) | 0.2 |

**Legend.** HFNC: high flow nasal cannula; NIV: non-invasive ventilation.

In **supplementary Table 1** is reported univariate analysis after propensity score matching to evaluate the impact of remdesivir-containing regimen on study population **(for reviewers’ information only). Figure 1** showed Kaplan Meier curves about 30-day survival in patients treated or not with remdesivir before (p=0.24) and after (p=0.07) propensity score matching, reporting no differences between the 2 study groups. Standardized differences before and after propensity score matching were reported in **supplementary Figure 1 (for reviewers’ information only)**.

**Figure 1.**
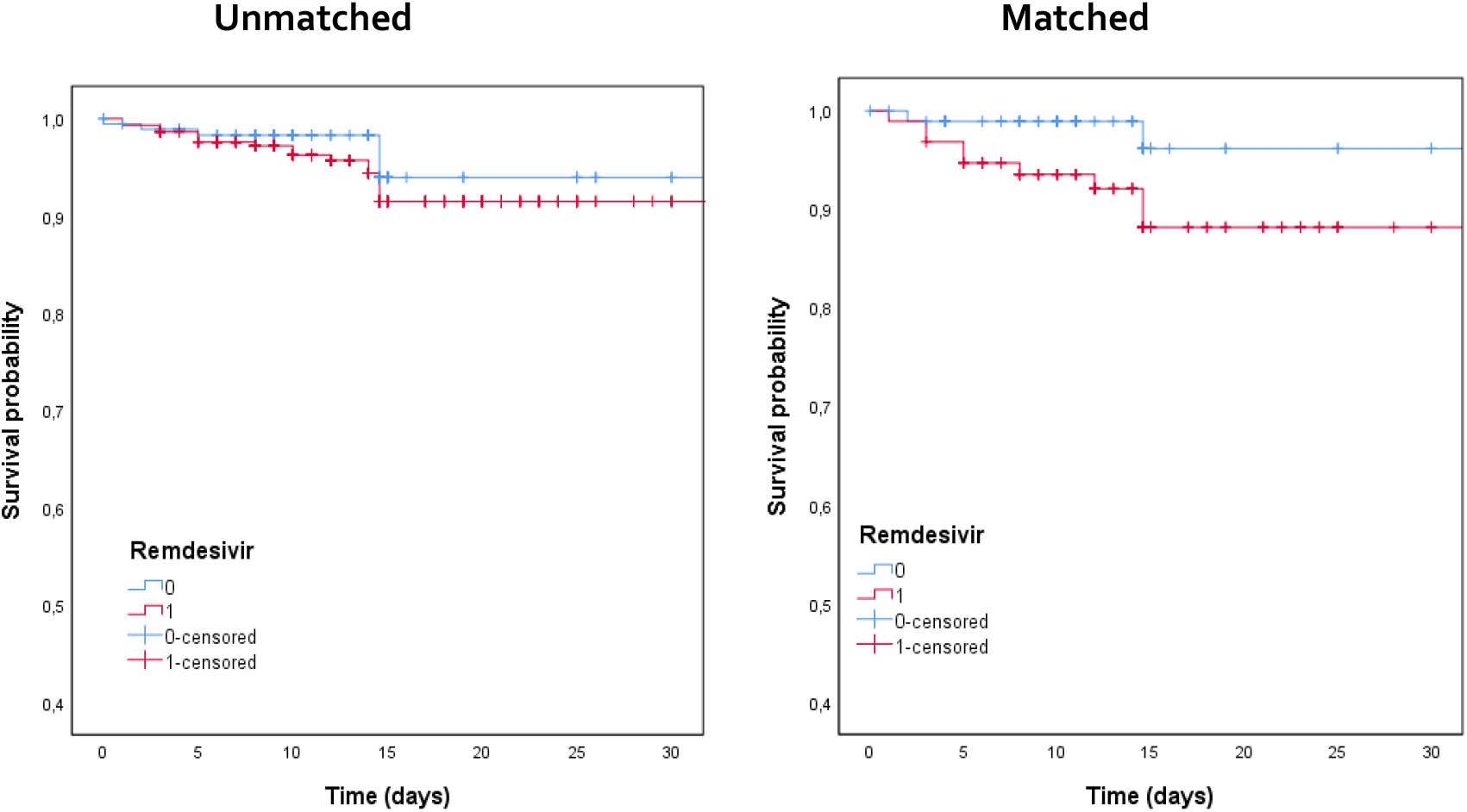
Kaplan Meier curves about 30-day survival in patients treated or not with remdesivir before (p=0.24) and after (p=0.07) propensity score matching

Multivariate Cox regression analysis about 30-day mortality after propensity score matching was reported in **Table 3**. Comorbidities and therapies, including remdesivir-containing therapy, were not statistically associated with 30-day survival or mortality. Instead, HFNC/NIV (HR 17.921, CI95% 0.954-336.73, p=0.044) and mechanical ventilation (HR 3.9, CI95% 5.36-16.2, p=0.003) resulted independently associated with 30-day mortality

**Table 3.** Multivariate Cox regression analysis about 30-day mortality after propensity score matching

| Variables | HR | CI95% | P value |
| --- | --- | --- | --- |
| Comorbidities | 0.136 | 0.009-2.04 | 0.149 |
| Chronic kidney disease | 1.773 | 0.022-140.18 | 0.797 |
| COPD | 3.033 | 0.097-95.03 | 0.528 |
| Bacterial co-infection | 0.693 | 0.061-7.81 | 0.767 |
| Low-molecular-weight heparin | 0.169 | 0.013-2.17 | 0.173 |
| Macrolides | 23.056 | 0.273-19,8 | 0.165 |
| Antibiotics (excluding macrolides) | 1.512 | 0.18-12.67 | 0.703 |
| Steroids | 0.1 | 0.0-1.2 | 0.89 |
| Low-flow oxygen therapy/no need of oxygen therapy | 11.682 | 0.575-237.45 | 0.11 |
| Remdesivir | 0.87 | 0.12-1.17 | 0.18 |
| HFNC/NIV | 17.921 | 0.954-336.73 | **0.044** |
| Mechanical ventilation | 3.9 | 5.36-16.2 | **0.003** |

**Legend.** COPD: chronic obstructive pulmonary disease; HFNC: high flow nasal cannula; NIV: non-invasive ventilation.

Finally, multivariate Cox regression analysis about need of non-invasive or invasive ventilation was analyzed after propensity score matching (see **Table 4**). Data showed that comorbidities and therapies, including remdesivir-containing regimen, were not independently associated with a lower or higher risk of HFNC/NIV or mechanical ventilation.

**Table 4.** Multivariate Cox regression analysis about need of non-invasive or invasive ventilation after propensity score matching

| Variables | HR | IC | P value |
| --- | --- | --- | --- |
| Comorbidities | 0.1 | 0.09-1.4 | 0.149 |
| Chronic kidney disease | 1.8 | 0.24-4.2 | 0.797 |
| COPD | 1.03 | 0.068-15.64 | 0.528 |
| Bacterial co-infection | 0.7 | 0.1-1.81 | 0.767 |
| Low-molecular-weight heparin | 0.169 | 0.01-2.17 | 0.173 |
| Macrolides | 0.31 | 0.01-8.8 | 0.493 |
| Antibiotics (excluding Macrolides) | 1.51 | 0.18-12.67 | 0.703 |
| Steroids | 3.2 | 0.78-136.9 | 0.890 |
| Remdesivir | 1.03 | 0.68-15.6 | 0.737 |

**Legend.** COPD: chronic obstructive pulmonary disease.

## DISCUSSION

This prospective clinical study reported a real-life experience about the use of remdesivir in a large population of consecutively hospitalized patients with COVID-19. Our data, also after propensity score matching, showed that remdesivir-containing regimen was not associated with 30-day survival compared to patients treated with other therapies not including remdesivir. Moreover, remdesivir-containing regimen was not independently related to progression or not to HFNC/NIV or mechanical ventilation.

In Italy, remdesivir was specifically licensed for the treatment of COVID-19 in hospitalized patients with pneumonia, requiring oxygen therapy but not treated with HFNC/NIV or mechanical ventilation at time of remdesivir prescription [9].

Different data were worldwide reported about the efficacy of remdesivir, taking in account different outcomes. Remdesivir treatment was associated with significantly higher recovery rates and lower mortality, compared to standard-of-care treatment without remdesivir in patients with severe COVID-19 [10]. In this study, significantly lower mortality was observed in those treated with remdesivir (7.6%) compared with the non-remdesivir-cohort patients (12.5%). Conversely, data from Solidarity trial, conducted in 30 countries [11], showed no decrease of in-hospital mortality in patients treated with remdesivir, with the important limitation that other outcomes, clinical improvement and adverse events, were non carefully evaluated.

Some important meta-analysis showed that COVID-19 patients receiving remdesivir had significantly higher rates of recovery and hospital discharge with lower rates of developing serious adverse events compared to patients receiving standard of care/placebo [12-13]. However, these analyses confirmed that no significant differences were observed about clinical improvement and rate of mortality during hospitalization. Specifically, mortality was the main outcome reported in all included studies, and none of the studies showed significant decrease of mortality also if they were not adequately powered for mortality outcome [12].

Wang et al. [14] reported the first double blinded, randomized, clinical trial in which were evaluated patients with an interval from symptoms onset to enrollment of 12 days or less. No differences in mortality were recorded in the 2 arms, also if authors highlighted a possible trend towards clinical benefit in remdesivir group. Of importance, a large number of patients in this study were treated also with steroids (65% of patients who received remdesivir and 68% of patients in placebo arm) which may have confounded the results and the conclusions. A strength of our study, with the limit of the non-randomized cohort, was to weight all the possible therapeutic confounders, comprising the use of steroids and LMWH.

Beigel et al. [8] randomized 1062 patients, hospitalized with COVID-19 and evidence of pneumonia, to remdesivir or placebo. The study demonstrated that remdesivir was superior to placebo in shortening the time to recovery in hospitalized COVID-19 patients with a trend towards survival benefit at day 29 without statistically significant differences. Of interest, authors reported a beneficial effect of remdesivir in severe COVID-19 patients but not requiring mechanical ventilation at enrollment; they suggested to start remdesivir early in the disease course.

Finally, in another randomized, clinical trial [15] in patients with moderate COVID-19 (no oxygen requirements, also if about 15% of patients required oxygen at time of enrollment) authors randomized 596 patients in 1:1:1 ratio to receive a 5-day course of remdesivir, a 10-day course of remdesivir, or standard of care therapy. In this study patients randomized to 5-day, but not 10-day treatment duration, showed a statistically significant difference in clinical status. A subgroup analysis excluding patients who required oxygen at baseline showed a significant difference favouring remdesivir over standard care.

Our study has some limitations. First, considering the monocentric design these results might be affected by local practice in the management of COVID-19; second, although criteria for HFNC/NIV or mechanical ventilation were based on the degree of respiratory impairment, elderly critically ill patients with ultimately fatal diseases were probably excluded from non-invasive/invasive ventilation, modifying the interpretation of some interventions. Second, in this analysis were evaluated all consecutively, hospitalized patients independently from COVID-19 severity, as demonstrated also by the low mortality (6% of remdesivir group Vs 4% of patients not treated with remdesivir). Finally, the analysis on the beneficial effects of treatments should be interpreted cautiously, because it was not conducted on randomized groups and might therefore be affected by several measured and unmeasured confounding factors. However, the comparison of remdesivir-treated patients with non-remdesivir-treated patients was based on a robust statistical methodology, appropriate for non-randomized cohort studies about therapy.

In conclusion, in our real-life experience about the remdesivir use in hospitalized patients with COVID-19 was not associated with significant increase in rates of survival or reduced use of HFNC/NIV or mechanical ventilation, compared to patients treated with other therapies not including remdesivir. These results suggest the need of more RCTs to evaluate the role of remdesivir in COVID-19 patients at different stages of disease or in combination with other drugs [16]. However, considering its safety profile and the lack of alternative drugs, remdesivir should be continued to be administered for patients with COVID-19.

## METHODS

### Study Design and Data Collection

This prospective, observational study included, from October 2020 to February 2021, patients admitted to University Hospital of Rome ‘‘Policlinico Umberto I‘’, in Italy. Inclusion criteria were: 1) positive SARS-CoV-2 real-time polymerase chain reaction test or an antigenic test on a nasopharyngeal swab; 2) pneumonia diagnosed either by CT thorax or chest x-ray; 3) need of hospitalization. Patients who required high flow oxygen therapy (HFNC) or non-invasive ventilation (NIV) or mechanical ventilation at time of hospitalization were excluded from this analysis.

All patients were evaluated in a dedicated Emergency Department by a dedicated staff of infectious diseases specialists that identified patients with SARS-CoV-2 pneumonia as soon as they arrived at the hospital, followed the patients during the hospital stay, and collected all data prospectively without interfering with the therapeutic decisions. This observational study was conducted according to the principles stated in the Declaration of Helsinki, and it conforms to standards currently applied in our country. The study was approved by the local EC. The patient’s informed consent was obtained.

Data were extracted from the medical records of patients and from hospital computerized databases. The following data were collected: demographics, clinical and laboratory findings, comorbidities, Charlson comorbidity index, microbiologic data, date of COVID-19 diagnosis, radiological characteristics of the pneumonia, therapies, concomitant infections, duration of mechanical ventilation, time of nasopharyngeal swab negativity, need of oxygen or ventilation support during the hospital stay, length of ICU stay, length of hospital stay. Development of moderate to severe ARDS was defined as the acute onset of hypoxemia, manifestations of pneumonia on chest computed tomography imaging of a noncardiac origin, and a PaO2/FiO2 ratio of less than 200 mmHg according to the Berlin Definition [17].

Remdesivir was administrated, after written informed consent, to patients with the following characteristics: pneumonia, less than 10 days from the onset of symptoms, no need for HFNC or NIV or mechanical ventilation, alanine aminotransferase no more than 5 times the upper limit of the reference range, estimated glomerular filtration rate (eGFR) greater than 30 mL/minute. A 5-day regimen was prescribed in all cases. Patients without these criteria were not eligible for remdesivir treatment.

All patients were followed-up until discharge or death.

## Endpoints and Statistical analysis

The primary endpoint of the study was to evaluate the impact of remdesivir-containing therapy on 30-day mortality in hospitalized patients with SARS-CoV2 pneumonia. Secondary endpoint was the impact of remdesivir-containing therapy on the need of NIV or mechanical ventilation.

To reduce the impact of treatment-selection bias in the estimation of treatment effects, propensity score matching was conducted. Variables were selected for inclusion in the propensity score based on potential impact on receipt of remdesivir and association with mortality [18]. The variables included were steroids, antibiotics (excluding macrolides), age, gender, and the use of LMWH during hospital admission. A propensity score density plot and Love plot were generated to examine the balance of propensity score and covariate distribution between the two groups (see **supplementary Figure 1)**.

To evaluate demographic factors, Welch’s t tests assuming unequal variances were used for continuous independent variables, while Pearson Chi-square or Fisher’s Exact Test when appropriate were used for categorical variables. Welch’s analysis of variance (ANOVA) was used to assess group differences for continuous outcome. Welch’s t-tests assuming unequal variances were used for post-hoc comparisons.

All tests will be two-tailed, and a p-value of <0.05 were considered statistically significant. Results were expressed as the mean with standard deviation (±SD) for continuous normally distributed variables and as a count (n) and percentage (%) for categorical variables. Multivariate analysis was used to identify independent predictors of 30-day mortality and need of NIV or mechanical ventilation. Matched bivariate analyses were conducted using a conditional logistic regression model, incorporating all variables found to be significant in the Univariate analysis (p < 0.05) with a stepwise method. Matched multivariate models was constructed using Cox proportional hazards (HRs) regression if appropriate, accounting for clustering on matched pairs. The final selected model was tested for confounding. In addition, 95% confidence intervals were calculated for HR. Survival was analyzed by Kaplan-Meier curves. All data were analyzed using a commercially available statistical software package (SPSS Statistics for Mac, 22.0; IBM Corp., Armonk, NY).

## Data Availability

Data are available on request to Prof. Alessandro Russo.

## Conflict of interest

none to declare.

## Funding

none.

## Transparency declarations

none to declare.

## Notes

### Competing Interest Statement

The authors have declared no competing interest.

### Funding Statement

No funding.

### Author Declarations

Sapienza University of Rome, local EC.

## REFERENCES

1 https://covid19.who.int, Accessed on 27 April 2021.

2 Russo A, Bellelli V, Ceccarelli G, Marincola Cattaneo F, Bianchi L, Pierro R, Russo R, Steffanina A, Pugliese F, Mastroianni CM, d’Ettorre G, Sabetta F. 2020. Comparison between hospitalized patients affected or not by COVID-19 (RESILIENCY study). Clin Infect Dis. 2020 Nov 18:ciaa1745. doi: 10.1093/cid/ciaa1745. Epub ahead of print.

3 Vistoli F, Furian L, Maggiore U, Caldara R, Cantaluppi V, Ferraresso M, Zaza G, Cardillo M, Biancofiore G, Menichetti F, Russo A, Turillazzi E, Di Paolo M, Grandaliano G, Boggi U; Italian National Kidney Transplantation Network; the Joint Committee of the Italian Society of Organ Transplantation and the Italian Society of Nephrology. 2020. COVID-19 and kidney transplantation: an Italian Survey and Consensus. J Nephrol. 33:667–680.

4 RECOVERY Collaborative Group, Horby P, Lim WS, Emberson JR, Mafham M, Bell JL, Linsell L, Staplin N, Brightling C, Ustianowski A, Elmahi E, Prudon B, Green C, Felton T, Chadwick D, Rege K, Fegan C, Chappell LC, Faust SN, Jaki T, Jeffery K, Montgomery A, Rowan K, Juszczak E, Baillie JK, Haynes R, Landray MJ. 2021. Dexamethasone in Hospitalized Patients with Covid-19. N Engl J Med. 384:693–704.

5 Falcone M, Tiseo G, Barbieri G, Galfo V, Russo A, Virdis A, Forfori F, Corradi F, Guarracino F, Carrozzi L, Celi A, Santini M, Monzani F, De Marco S, Pistello M, Danesi R, Ghiadoni L, Farcomeni A, Menichetti F; Pisa COVID-19 Study Group. 2020. Role of Low-Molecular-Weight Heparin in Hospitalized Patients With Severe Acute Respiratory Syndrome Coronavirus 2 Pneumonia: A Prospective Observational Study. Open Forum Infect Dis. 7:ofaa563

6 Wang M, Cao R, Zhang L, Yang X, Liu J, Xu M, Shi Z, Hu Z, Zhong W, Xiao G. 2020. Remdesivir and chloroquine effectively inhibit the recently emerged novel coronavirus (2019-nCoV) in vitro. Cell Res. 30:269–271.

7 Frediansyah A, Nainu F, Dhama K, Mudatsir M, Harapan H. Remdesivir and its antiviral activity against COVID-19: A systematic review. Clin Epidemiol Glob Health. 2021;9:123–127.

8 Beigel JH, Tomashek KM, Dodd LE, Mehta AK, Zingman BS, Kalil AC, Hohmann E, Chu HY, Luetkemeyer A, Kline S, Lopez de Castilla D, Finberg RW, Dierberg K, Tapson V, Hsieh L, Patterson TF, Paredes R, Sweeney DA, Short WR, Touloumi G, Lye DC, Ohmagari N, Oh MD, Ruiz-Palacios GM, Benfield T, Fätkenheuer G, Kortepeter MG, Atmar RL, Creech CB, Lundgren J, Babiker AG, Pett S, Neaton JD, Burgess TH, Bonnett T, Green M, Makowski M, Osinusi A, Nayak S, Lane HC; ACTT-1 Study Group Members. 2020. Remdesivir for the Treatment of Covid-19 - Final Report. N Engl J Med. 383:1813–1826.

9 https://www.aifa.gov.it/-/procedura-di-richiesta-per-il-farmaco-veklury-remdesivir. Accessed on 28 April 2021.

10 Olender SA, Perez KK, Go AS, Balani B, Price-Haywood EG, Shah NS, Wang S, Walunas TL, Swaminathan S, Slim J, Chin B, De Wit S, Ali SM, Soriano Viladomiu A, Robinson P, Gottlieb RL, Tsang TYO, Lee IH, Haubrich RH, Chokkalingam AP, Lin L, Zhong L, Bekele BN, Mera-Giler R, Gallant J, Smith LE, Osinusi AO, Brainard DM, Hu H, Phulpin C, Edgar H, Diaz-Cuervo H, Bernardino JI. 2020. Remdesivir for Severe COVID-19 versus a Cohort Receiving Standard of Care. Clin Infect Dis. 2020 Jul 24:ciaa1041. doi: 10.1093/cid/ciaa1041. Epub ahead of print.

11 WHO Solidarity Trial Consortium, Pan H, Peto R, Henao-Restrepo AM, Preziosi MP, Sathiyamoorthy V, Abdool Karim Q, Alejandria MM, Hernández García C, Kieny MP, Malekzadeh R, Murthy S, Reddy KS, Roses Periago M, Abi Hanna P, Ader F, Al-Bader AM, Alhasawi A, Allum E, Alotaibi A, Alvarez-Moreno CA, Appadoo S, Asiri A, Aukrust P, Barratt-Due A, Bellani S, Branca M, Cappel-Porter HBC, Cerrato N, Chow TS, Como N, Eustace J, García PJ, Godbole S, Gotuzzo E, Griskevicius L, Hamra R, Hassan M, Hassany M, Hutton D, Irmansyah I, Jancoriene L, Kirwan J, Kumar S, Lennon P, Lopardo G, Lydon P, Magrini N, Maguire T, Manevska S, Manuel O, McGinty S, Medina MT, Mesa Rubio ML, Miranda-Montoya MC, Nel J, Nunes EP, Perola M, Portolés A, Rasmin MR, Raza A, Rees H, Reges PPS, Rogers CA, Salami K, Salvadori MI, Sinani N, Sterne JAC, Stevanovikj M, Tacconelli E, Tikkinen KAO, Trelle S, Zaid H, Røttingen JA, Swaminathan S. 2021. Repurposed Antiviral Drugs for Covid-19 - Interim WHO Solidarity Trial Results. N Engl J Med. 384:497–511.

12 Al-Abdouh A, Bizanti A, Barbarawi M, Jabri A, Kumar A, Fashanu OE, Khan SU, Zhao D, Antar AAR, Michos ED. 2021. Remdesivir for the treatment of COVID-19: A systematic review and meta-analysis of randomized controlled trials. Contemp Clin Trials. 101:106272.

13 Wilt TJ, Kaka AS, MacDonald R, Greer N, Obley A, Duan-Porter W. 2021. Remdesivir for Adults With COVID-19 : A Living Systematic Review for American College of Physicians Practice Points. Ann Intern Med. 174:209–220.

14 Wang Y, Zhang D, Du G, Du R, Zhao J, Jin Y, Fu S, Gao L, Cheng Z, Lu Q, Hu Y, Luo G, Wang K, Lu Y, Li H, Wang S, Ruan S, Yang C, Mei C, Wang Y, Ding D, Wu F, Tang X, Ye X, Ye Y, Liu B, Yang J, Yin W, Wang A, Fan G, Zhou F, Liu Z, Gu X, Xu J, Shang L, Zhang Y, Cao L, Guo T, Wan Y, Qin H, Jiang Y, Jaki T, Hayden FG, Horby PW, Cao B, Wang C. 2020. Remdesivir in adults with severe COVID-19: a randomised, double-blind, placebo-controlled, multicentre trial. Lancet. 395:1569-1578. Erratum in: Lancet. 395:1694.

15 Spinner CD, Gottlieb RL, Criner GJ, Arribas López JR, Cattelan AM, Soriano Viladomiu A, Ogbuagu O, Malhotra P, Mullane KM, Castagna A, Chai LYA, Roestenberg M, Tsang OTY, Bernasconi E, Le Turnier P, Chang SC, SenGupta D, Hyland RH, Osinusi AO, Cao H, Blair C, Wang H, Gaggar A, Brainard DM, McPhail MJ, Bhagani S, Ahn MY, Sanyal AJ, Huhn G, Marty FM; GS-US-540-5774 Investigators. 2020. Effect of Remdesivir vs Standard Care on Clinical Status at 11 Days in Patients With Moderate COVID-19: A Randomized Clinical Trial. JAMA. 324:1048–1057.

16 Kalil AC, Patterson TF, Mehta AK, Tomashek KM, Wolfe CR, Ghazaryan V, Marconi VC, Ruiz-Palacios GM, Hsieh L, Kline S, Tapson V, Iovine NM, Jain MK, Sweeney DA, El Sahly HM, Branche AR, Regalado Pineda J, Lye DC, Sandkovsky U, Luetkemeyer AF, Cohen SH, Finberg RW, Jackson PEH, Taiwo B, Paules CI, Arguinchona H, Erdmann N, Ahuja N, Frank M, Oh MD, Kim ES, Tan SY, Mularski RA, Nielsen H, Ponce PO, Taylor BS, Larson L, Rouphael NG, Saklawi Y, Cantos VD, Ko ER, Engemann JJ, Amin AN, Watanabe M, Billings J, Elie MC, Davey RT, Burgess TH, Ferreira J, Green M, Makowski M, Cardoso A, de Bono S, Bonnett T, Proschan M, Deye GA, Dempsey W, Nayak SU, Dodd LE, Beigel JH; ACTT-2 Study Group Members. 2021. Baricitinib plus Remdesivir for Hospitalized Adults with Covid-19. N Engl J Med. 384:795–807.

17 ARDS Definition Task Force, Ranieri VM, Rubenfeld GD, Thompson BT, Ferguson ND, Caldwell E, Fan E, Camporota L, Slutsky AS. Acute respiratory distress syndrome: the Berlin Definition. 2012. JAMA. 307:2526–33.

18 Rajendram P, Sacha GL, Mehkri O, Wang X, Han X, Vachharajani V, Duggal A. 2021. Tocilizumab in Coronavirus Disease 2019-Related Critical Illness: A Propensity Matched Analysis. Crit Care Explor. 3:e0327

